# Choice of Intraoperative Ultrasound adjuncts for Brain Tumor Surgery

**DOI:** 10.1101/2022.02.02.22270289

**Authors:** Manoj Kumar, Santosh Noronha, Narayan Rangaraj, Aliasgar Moiyadi, Prakash Shetty, Vikas Kumar Singh

## Abstract

**Background:** Gliomas are among the most typical brain tumors tackled by neurosurgeons. During navigation for surgery of glioma brain tumors, preoperatively acquired static images may not be accurate due to shifts. Surgeons use intraoperative imaging technologies (2-Dimensional and navigated 3 Dimensional ultrasound) to assess and guide resections. This paper aims to precisely capture the importance of preoperative parameters to decide which type of ultrasound to be used for a particular surgery.

**Methods:** This paper proposes two bagging algorithms considering base classifier logistic regression and random forest. These algorithms are trained on different subsets of the original data set. The goodness of fit of Logistic regression-based bagging algorithms is established using hypothesis testing. Furthermore, the performance measures for random-forest-based bagging algorithms used are AUC under ROC and AUC under the precision-recall curve. We also present a composite model without compromising the explainability of the models.

**Results:** These models were trained on the data of 350 patients who have undergone brain surgery from 2015 to 2020. The hypothesis test shows that a single parameter is sufficient instead of all three dimensions related to the tumor (*p* < 0.05). We observed that the choice of intraoperative ultrasound depends on the surgeon making a choice, and years of experience of the surgeon could be a surrogate for this dependence.

**Conclusion:** This study suggests that neurosurgeons may not need to focus on a large set of preoperative parameters in order to decide on ultrasound. Moreover, it personalizes the use of a particular ultrasound option in surgery. This approach could potentially lead to better resource management and help healthcare institutions improve their decisions to make the surgery more effective.

## Background

Gliomas are among the commonest brain tumors encountered by neurosurgeons. Surgery is an integral component of its treatment, and the extent of resection is a crucial prognostic factor. Due to the ill-defined nature of these tumors, surgeons increasingly rely on technological adjuncts to identify and remove maximum tumor safely. Navigation or frame-less stereotaxy is the standard tool based on preoperatively acquired MR images which acts like a GPS providing a road map for the surgical procedure. However, due to the changes in anatomical structures during surgery, its accuracy is compromised, necessitating updated intraoperative imaging.

Tumor surgery has two main stages - lesion localization, and control of resection during surgery [1]. State-of-the-art intraoperative micro-surgical techniques are supplemented by techniques such as Magnetic Resonance Imaging (MRI), CT, and ultrasonography to improve the real-time updates [2].

Although intraoperative MRI (iMRI) would be ideal, it is not widely available and is costly. Intraoperative ultrasound (US) has emerged as a very popular solution [3], both 2D US (standard default US modality) as well as advanced navigated 3D US (3DUS). 3DUS provides navigated multiplanar images often with fusion imaging with preoperative MR and is believed to provide better orientation and image interpretation, thereby making it a viable option.

However, the navigated 3D ultrasound costs more as one scan takes 3 to 5 minutes (1 to 2 minutes for 2D), and the setup cost is higher than 2D ultrasound. Thus there is a trade-off between these imaging technologies, and one of the objectives in this paper is to analyze how neurosurgeons choose to deploy these two modalities during routine practice.

Different studies have shown that the resolution of ultrasound images deteriorates as the surgery progresses [1, 4]. Thus, navigated 3D ultrasound might not be beneficial in later stages.

In this study, we attempt to understand the preoperative factors that affect the choice of ultrasound.

### Objective of the study

This study proposes a data-driven optimal decision policy based on patients and tumor characteristics. We have tried to answer the following questions:

1. What attributes of the patient and the tumor affect the choice of ultrasound?
2. Does the experience of the surgeon affect the decision?

We have used a real-world surgery data set gathered from a tertiary care referral neurosurgical oncology centre. It has the preoperative MRI tumor information and patient characteristics of 350 patients. We have used random forest and logistic regression models to analyze this data.

While assessing the benefits of certain technical adjuncts in healthcare, it is important to understand the patterns of use during routine care, which may differ from those under controlled trial conditions. Routine practices reflect day-to-day factors which are often difficult to pinpoint in preliminary observation. These factors need to be better understood to make conscious and well-informed decisions regarding the deployment of such health care technologies. This is more important if the eventual outcomes are affected by this choice or if there is a significant cost-consequence of these choices. In health care situations, it is often very difficult to test the effect of different states of the same factor (different types of techniques/ adjuncts) due to practical and logistical difficulties. Using large databases and employing rigorous data science methods may be the best option.

The major conclusion is that surgeon’s experience, prior treatment, and contrast enhancement pattern variables are statistically significant in most models. The patient’s age is the only demographic factor that is statistically significant.

The rest of the paper is organized as follows. In section, we presented the comprehensive literature survey related to glioma tumor surgery and various techniques used in medical decision-making. We have elaborated the problem and data description in section and presented the statistical analysis results in section. Section details the methodology used in this study. Data analysis results are provided in section. Section elaborates the discussion and section states the conclusion of the paper.

### Literature Survey

#### Surgical Workflow Analysis

The use of adjuncts needs to be understood in the context of surgical workflow. This workflow has different components, including low-level tasks, high-level tasks, patient status, and the use of medical devices. A study has been proposed to classify these situations based on multi-perceptive analysis [5]. Medical devices are developed stand-alone to provide specific functionality for a certain stage of the surgery. In [6], the authors have presented a model-driven design of surgical workflow to map the information of all these devices.

Surgeons need to make decisions about various tasks during surgical operations, called intraoperative decisions. Different situations and strategies in general are discussed in [7].

#### Glioma Surgery

An automatic estimation method for brain tumor resection was developed in [8] based on the anatomical information received by the surgical navigation system using a Bayesian technique. The surgical navigation systems’ stand-alone use fails to improve the outcome of brain tumor surgeries.

In the literature, many studies highlight the impact of intraoperative ultrasound for controlling the extent of resection of tumor tissue, for example [9, 3, 10]. A study [11] has been conducted to understand the applications and interaction between different modes of intraoperative imaging under the subjective basis of 11 surgical case studies. It highlights that iMRI is always the surgeon’s choice, while it is evident from the study that the beneficial imaging modality is linear array intraoperative ultrasound.

The authors in [12] showed that the superimposition of navigable 3D ultrasound with preoperative MRI provides a better orientation of the cross-sectional anatomy. Another study [1] showed that navigated 3D ultrasound without the preoperative images eliminates the registration inaccuracy inherent to image-to-patient registration algorithms. Another study has compared image-guided surgery with surgery being performed by either not using any image guidance or using two different forms of image guidance [2].

#### Statistical Analysis in Healthcare

In medical decision-making, different statistical techniques have been widely used to improve the understanding of medical practitioners. Logistic regression [13] is widely used in healthcare to find the relationship between different parameters. Other techniques used to improve model performance are principal component analysis [14, 15] and bootstrap sampling [13].

#### Machine learning in healthcare

Many researchers have used machine learning algorithms in a variety of applications like the diagnosis of heart failure [16], diabetes [17], analysis of MRI data to quantify the neurological disorder [18], predicting diabetic retinopathy [19], and tumor segmentation [17]. Decision support systems have been proposed using the predictive models of data mining for overall survivability with comorbidity, of cancers [20] and graft survival among kidney transplant recipients [21]. The receiver operating characteristic curve (ROC) [22] is popularly used as the measure of performance for many machine learning algorithms. Convolutional neural networks have been proposed to diagnose gastric endoscopy-based gastric cancer, and they performed better than human pathologists [23].

A random forest and Cox proportional-hazard model has been developed to assess the association between contrast enhancement pattern of IHD mutant and diffuse glioma tumor with survival [24].

While several studies have been done to understand the parts of brain cancer workflow and the adjuncts used during surgery, benefits discussed in these studies are stand-alone and do not include the decision regarding the imaging modality to use in a particular patient case. Most importantly, none address the factors that influence the choice of using a particular technical modality. Our study integrating the above aspects would be novel and relevant for the field of brain cancer surgery. This could better inform neurosurgeons on selecting the most suitable modality a priori and potentially dictate decision making when identifying and inducting appropriate adjuncts in setting up a service.

## Methods

### Problem & Data Description

Intraoperative imaging technologies play a vital role in brain cancer surgery. Some of the possible technologies are intraoperative 2-Dimensional ultrasound (2DUS), navigated 3-Dimensional ultrasound (3DUS), and Magnetic Resonance Imaging (MRI). We try to identify factors that govern the choice of using the different types of intraoperative ultrasound based on the demographic factors of the patient, surgeon’s experience, and tumor characteristics. These factors are known a priori and can be built into a decision-making algorithm during the preoperative stage allowing optimal allocation and utilization of resources as well as serving as a recommendation in different types of scenarios.

We also explore whether the surgeon’s personal choice affects intraoperative 2DUS vs. 3DUS decisions.

The data used in this analysis is secondary data collected from the electronic records of a tertiary care referral neurosurgical oncology centre. All patients undergoing resection for gliomas where intraoperative ultrasound was utilized and had preoperative MRI available for review during the time period 2015-2020 were analyzed. The use of anonymised retrospective data was approved for this study. Clinical and radiological features based on preoperative routine MRI were extracted. The attributes of interest included patient’s age, gender, prior treatment status (yes/ no), eloquent location (yes/ no), depth of tumor (surfacing/ sub-cortical/ deep), histology (high grade/ low grade), glioma location (lobar/ no-lobar), delineation (good/ moderate/ poor), contrast enhancement pattern (negligible/ mixed/ predominant), tumor dimensions in three orthogonal planes (height, length, width), and surgeon experience. Additionally, a variable spherical diameter was computed using the volume equivalent spherical diameter using MRI height, length, and width of the tumor.

We have included 350 procedures, out of which 2D ultrasound was used for 143 surgeries. Out of these three values were missing for contrast enhancement patterns, these were imputed using mode value.

In this data set, four surgeons have performed all the surgeries. The number of surgeries accomplished by a surgeon is taken as the surgeon’s experience. The average (St. Dev.) age of patients is 41.23 (14.71) years. Appropriate correlation methods measuring the association between the variables were applied [25] and are shown in Appendix A.

The correlation among the tumor’s length, height, and width is significant, and all other variables showed negligible correlation.

### Statistical Analysis

We have performed both parametric (t-test) and non-parametric (Mann-Whitney test) tests on the data sets to confirm the normality of the data. We have presented the p-values corresponding to the Mann-Whitney test here as we obtained the same result from both methods. The hypothesis tested is that both groups of technologies result in the same mean/ proportions for the variables listed.

Surgeon experience and prior treatment status are statistically significant in both groups. The most experienced surgeon has used the navigated 3D ultrasound more often. The average age of the patients, length, and height are more in 3DUS group but not statistically significant.

The data set is stratified into two groups - surgeon group 1, which includes the patients whose surgery was performed by the most experienced surgeon, and surgeon group 2 that consists of the patients whose surgeries were performed by three other surgeons.

Surgeon group 1 has performed 214 surgeries, out of which 58 (27%) surgeries are with 2DUS. Table 2 depicts the description of surgeon group 1 data set. The t-test and Mann-Whitney tests showed that none of the attributes are statistically significant.

**Table 1:** Description of complete data set. * denotes *p* < 0.05, p-value are corresponding to Mann-Whitney test.

|  |  | 2D (n = 143) | 3DUS (n= 207) | p-value |
| --- | --- | --- | --- | --- |
|  |  | Mean (St. Dev.) |  |  |
| Age (in years) |  | 40.32 (15.72) | 41.86 (13.97) | 0.35 |
| Length (in cm) |  | 4.81 (1.59) | 4.98 (1.65) | 0.21 |
| Width (in cm) |  | 3.91 (1.18) | 3.83 (1.14) | 0.53 |
| Height (in cm) |  | 4.03 (1.30) | 4.15 (1.33) | 0.34 |
| Surgeon Experience (no. of surgeries) |  | 143.17 (64.65) | 184.23 (55.20) | 0.00* |
| Gender | Male | 97 | 149 | 0.4 |
|  | Female | 46 | 58 |  |
| Prior Treatment | Yes | 45 | 36 | 0.00* |
|  | No | 98 | 171 |  |
| Eloquent Location | Yes | 55 | 98 | 0.1 |
|  | No | 88 | 109 |  |
| Depth of Tumor | Surfacing | 68 | 113 | 0.26 |
|  | Sub-cortical | 41 | 49 |  |
|  | Deep | 34 | 45 |  |
| Histology | Low Grade | 31 | 49 | 0.66 |
|  | High Grade | 112 | 158 |  |
| Glioma Location | Lobar | 135 | 203 | 0.06 |
|  | No-Lobar | 8 | 4 |  |
| Delineation | Poor | 10 | 23 | 0.97 |
|  | Moderate | 80 | 102 |  |
|  | Good | 53 | 82 |  |
| Contrast Enhancement Pattern | Negligible | 33 | 78 | 0.30 |
|  | Mixed | 88 | 80 |  |
|  | Predominant | 22 | 49 |  |

**Table 2:** Description of surgeon group 1 data. p-values are corresponding to MannWhitney test.

|  |  | 2D (n = 58) | 3DUS (n= 156) | p-value |
| --- | --- | --- | --- | --- |
|  |  | Mean(st. dev.) |  |  |
| Age (in years) |  | 43.79 (16.64) | 41.42 (13.77) | 0.43 |
| Gender | Male | 39 | 114 | 0.4 |
|  | Female | 19 | 42 |  |
| Prior Treatment | Yes | 17 | 27 | 0.05 |
|  | No | 41 | 129 |  |
| Eloquent Location | Yes | 25 | 79 | 0.32 |
|  | No | 33 | 77 |  |
| Depth of Tumor | Surfacing | 26 | 88 | 0.18 |
|  | Sub-cortical | 18 | 36 |  |
|  | Deep | 14 | 32 |  |
| Histology | Low Grade | 12 | 34 | 0.86 |
|  | High Grade | 46 | 122 |  |
| Glioma Location | Lobar | 54 | 153 | 0.07 |
|  | No-Lobar | 4 | 3 |  |
| Delineation | Poor | 3 | 16 | 0.59 |
|  | Moderate | 32 | 80 |  |
|  | Good | 23 | 60 |  |
| Contrast Enhancement Pattern | Negligible | 12 | 63 | 0.22 |
|  | Mixed | 36 | 54 |  |
|  | Predominant | 10 | 39 |  |

Surgeon group 2 has performed 136 surgeries, out of which 51 (37.5%) surgeries are with navigated 3D ultrasound. Mann-Whitney test shows that none of the parameters (taken one at a time) are statistically significant except prior treatment, which is at borderline, as shown in Table 3.

**Table 3:** Description of surgeons group 2 data set. p-values are corresponding to Mann-Whitney test.

|  |  | 2D (n = 85) | 3DUS (n= 51) | p-value† |
| --- | --- | --- | --- | --- |
|  |  | Mean(St. Dev.) |  |  |
| Age (in years) |  | 37.95 (14.70) | 43.23 (14.64) | 0.07 |
| Gender | Male | 58 | 35 | 0.96 |
|  | Female | 27 | 16 |  |
| Prior Treatment | Yes | 28 | 9 | 0.05 |
|  | No | 57 | 42 |  |
| Eloquent Location | Yes | 30 | 19 | 0.82 |
|  | No | 55 | 32 |  |
| Depth of Tumor | Surfacing | 42 | 25 | 0.89 |
|  | Sub-cortical | 23 | 13 |  |
|  | Deep | 20 | 13 |  |
| Histology | Low Grade | 19 | 15 | 0.36 |
|  | High Grade | 66 | 36 |  |
| Glioma Location | Lobar | 81 | 50 | 0.41 |
|  | No-Lobar | 4 | 1 |  |
| Delineation | Poor | 7 | 7 | 0.69 |
|  | Moderate | 48 | 22 |  |
|  | Good | 30 | 22 |  |
| Contrast Enhancement Pattern | Negligible | 21 | 15 | 0.99 |
|  | Mixed | 52 | 26 |  |
|  | Predominant | 12 | 10 |  |

Thus, the major difference between surgeon groups 1 and 2 is that the former had more navigated 3D procedures, and the latter had more 2D ultrasound ones.

### Methodology

We have designed two bootstrap cum aggregation (bagging) algorithms with logistic regression, and random forest as base (weak) classifiers [26, 27]. The non-parametric bootstrap sampling technique [13] was used for generating different learning set from the data set. The bagging algorithms are an aggregation of weak classifiers trained on bootstrap samples. We aggregated the final prediction by averaging the predicted probabilities of each of the weak classifiers.

#### Data Analysis

Both bagging algorithms have been trained on the complete data set, on various subsets, and after dimensional reduction of the data set. We have also combined some levels of ordinal features and trained the logistic regression and random forest classifier to develop a composite model. All these data sets were standardized to standard normal distribution beforehand. All the data sets are divided randomly in the training set (80%) and testing test (20%), and models were trained on bootstraps samples drawn from training data sets.

#### Actual data set analysis

In this section, we have discussed the models trained on the actual data set and their corresponding results.

#### Complete data set

The complete data set was used to generate 11000 bootstrap samples and logistic regression (referred as **LR-full model**) and random forest-based bagging (referred as **RF-full model**) were trained.

Odds ratio (OR) of patient’s gender, surgeon’s experience, eloquent location, length, height, and glioma location are greater than one, and for all other variables, they are less than one. OR > 1 indicates the likelihood of navigated 3D is higher as compared to 2D ultrasound, and OR < 1 indicates a decrease in the likelihood of navigated 3D ultrasound.

#### Complete data set with spherical diameter

In this data set, instead of height, length, and width, we have introduced a new parameter ‘spherical diameter’ which is computed from equivalent spherical volume 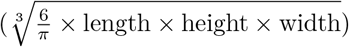. The logistic regression (**LR-Spherical model**) and random forest (**RF-Spherical model**) based bagging algorithm were trained on this dataset.

The spherical diameter variable is constructed because there is a high correlation between the tumor’s length, height, and width (tumors are relatively unlikely to grow along only one dimension). A *χ*^2^-test shows that the hypothesis that the spherical diameter is sufficient to capture the information of the three-dimension-related variables cannot be ruled out (*p* = 0.763 > 0.05). This allows us to make the models more compact.

The surgeon experience variable is a most important feature in RF-full and RF-Spherical shown in Figure (1) and has a larger coefficient in LR-full and LR-Spherical model as well. Also, its coefficient is statistically significant in both the models defined earlier. We have performed the analysis after excluding it to claim that this is indeed an important factor.

**Figure 1:**
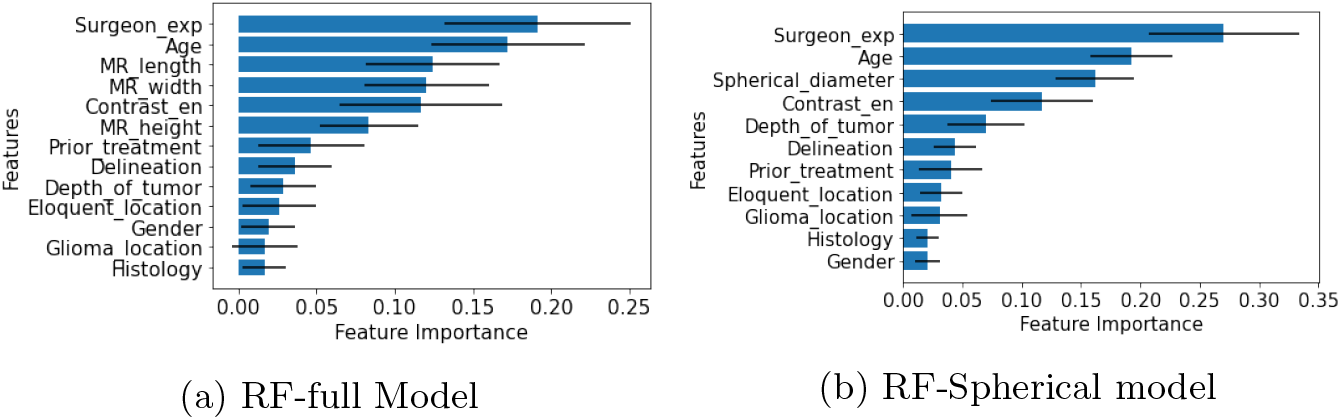
Feature Importance for random forest model: the bars show the 95% confidence interval centered on the mean value. Surgeon exp represents surgeon experience and contrast en represents contrast enhancement pattern.

#### Data set with surgeon experience removed

We have removed the surgeon’s experience from the complete data set and randomly divided the data set into training (80%) and testing set (20%). The bootstrap samples were drawn from the training set. The random forest (**RF-Surgeon’s Experience Removed**) and logistic regression-based bagging algorithm (**LR-Surgeon’s Experience Removed**) were trained on each of these bootstrap samples.

We have observed that the performance of both bagging algorithms worsened after dropping the surgeon experience feature.

The *χ*^2^-test showed that LR-Spherical model and LR-Surgeon’s Experience Removed are statistically different (*p* = 0.005 < 0.05). Therefore, removing the surgeon’s experience from the model increases the deviance of the model and thus degrades the performance of the model. Hence surgeon’s experience is an important factor in the choice of intraoperative ultrasound.

#### Surgeon based stratification

The complete data set is stratified into two groups based on the surgeons who have executed the surgeries. The details are discussed in the Table (2) and (3).

Surgeon group 1 has 27% 2DUS samples, and surgeon group 2 has 37.5% 3DUS samples. Class imbalance is when one class has more elements than another in the data set, which biases predictive models towards the majority class. To prevent this, we have used the Synthetic Minority Oversampling Technique (SMOTE) [28]. SMOTE over-samples the minority class using *k* nearest neighbors technique.

In surgeon group 1, we have considered the value of *k* as six. The balanced data set of surgeon group 1 was divided into training and test set with an 80:20 ratio. The logistic regression (**LR-Surgeon 1 group**) and random forest-based bagging algorithm (**RF-Surgeon 1 group**) were trained on 11500 bootstrap samples from the training set.

In surgeon group 2, we have over-sampled the navigated 3D ultrasound class using *k* is equal to 3 and divided the data into 80:20 ratio for training and testing set. The logistic regression (**LR-Surgeon 2 group**) and random forest-based bagging algorithm (**RF-Surgeon 2 group**) were trained on 11500 bootstrap samples of size equal to the training set.

The prior treatment is statistically significant in LR-full (*p* = 0.008), LR-Surgeon’s Experience Removed model (*p* = 0.023), LR-Surgeon 1 group model (*p* = 0.007), and at border line in LR-Spherical model. The width is statistically significant in LR-full model (*p* = 0.04). The LR-Surgeon 1 showed that patient’s age is statistically significant as *p* = 0.011, whereas histology (*p* = 0.018) in LR-Surgeon 2 model. Table (4) shows the likelihood of choice of ultrasound-based on the odds ratios corresponding to all logistic regression models discussed so far.

**Table 4:**
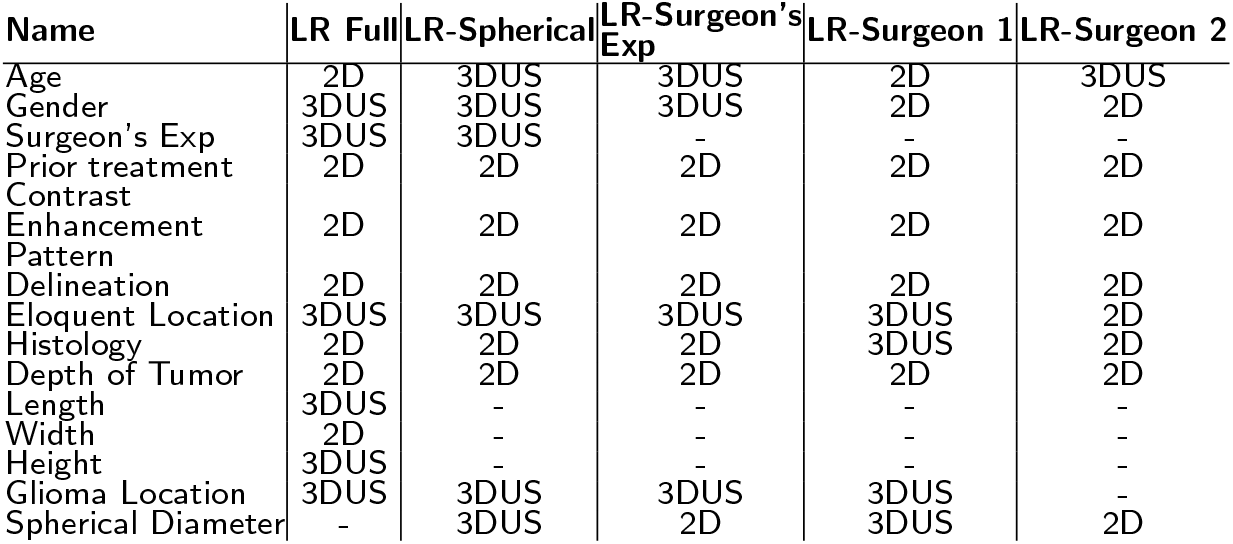
Likelihood of ultrasound of different logistic regression based bagging models. 2D represents the 2D ultrasound whereas 3DUS represents the navigated 3D ultrasound. - represents the exclusion of that variable in the model.

| Name | LR Full | LR-Spherical | LR-Surgeon's Exp | LR-Surgeon 1 | LR-Surgeon 2 |
| --- | --- | --- | --- | --- | --- |
| Age | 2D | 3DUS | 3DUS | 2D | 3DUS |
| Gender | 3DUS | 3DUS | 3DUS | 2D | 2D |
| Surgeon's Exp | 3DUS | 3DUS | - | - | - |
| Prior treatment | 2D | 2D | 2D | 2D | 2D |
| Contrast Enhancement Pattern | 2D | 2D | 2D | 2D | 2D |
| Delineation | 2D | 2D | 2D | 2D | 2D |
| Eloquent Location | 3DUS | 3DUS | 3DUS | 3DUS | 2D |
| Histology | 2D | 2D | 2D | 3DUS | 2D |
| Depth of Tumor | 2D | 2D | 2D | 2D | 2D |
| Length | 3DUS | - | - | - | - |
| Width | 2D | - | - | - | - |
| Height | 3DUS | - | - | - | - |
| Glioma Location | 3DUS | 3DUS | 3DUS | 3DUS | - |
| Spherical Diameter | - | 3DUS | 2D | 3DUS | 2D |

#### Analysis after redesigning of some parameters

The above models include the patient as well as tumor characteristics. It is very unlikely that patient’s age and gender would influence the choice of the US being used. Therefore, we will consider only the tumor characteristics visible to surgeons before starting the surgery in further models, but after redefining some of them, such as contrast enhancement pattern, delineation, and location of tumor.

The contrast enhancement pattern is redefined as follows: ‘predominant’ + ‘mixed’ is taken as enhancing, and ‘negligible’ is taken as ‘non-enhancing’.

Delineation is redefined as a dichotomous variable in two distinct ways:

1. Moderate grouped with poor delineation, and good delineation kept separate (denoted as PMD)
2. Moderate grouped with good, and poor delineation kept separate (denoted as GMD)

This is because the definition of moderate may be subjective, whereas poor and good delineations are more easily and reproducibly defined. The location of tumor was also redefined by coupling the depth of the tumor and height of the tumor as follows:

1. We club ‘surfacing’ and ‘sub-cortical’ tumors as ‘surfacing’ and assign all of them as fixed ‘surface depth’ value of 0.5 cm as these were defined as less than 1 cm. The ‘surface depth’ of deep tumors is considered as 1 cm.
2. Then we use the height value of each tumor, take its midpoint and add it to ‘surface depth’ (which is 0.5 or 1) to get the epicenter depth of the tumor.
3. Then we have defined the new variable location of tumor as ‘superficial’ if epicenter depth is less than 3 cm and otherwise ‘deep’ tumor.

A threshold of 3 cm is reasonable as total depth of the brain practically is observed to be 5 cm to 6 cm. The statistical analysis of all these variables is shown in Appendix B.

Based on this, we have constructed the following data set after suitable changes.

A. **Complete data set with PMD**: The logistic regression and random forest models trained on this data set are referred as **LR-PMD Spherical** and RF-PMD Spherical.
B. **Surgeon Stratified data sets with PMD**: The logistic regression and random forest models trained on these data sets are referred as **LR-PMD Spherical Surgeon 1, LR-PMD Spherical Surgeon 2, RF-PMD Spherical Surgeon 1**, and **RF-PMD Spherical Surgeon 2**.
C. **Complete data set with GMD**: The logistic regression and random forest models trained on this data set are referred as **LR-GMD Spherical** and **RF-GMD Spherical**.
D. **Surgeon stratified data sets with GMD**: The logistic regression and random forest models trained on these data sets are referred as **LR-GMD Spherical Surgeon 1, LR-GMD Spherical Surgeon 2, RF-GMD Spherical Surgeon 1**, and **RF-GMD Spherical Surgeon 2**.

In (A) and (B), we have only included the tumor characteristics with delineation defined as PMD, whereas in (C) and (D), delineation is defined as GMD. We have trained different logistic regression and random forest models on these data sets. Table (5) shows the likelihood of choice of intraoperative ultrasound in different models.

**Table 5:**
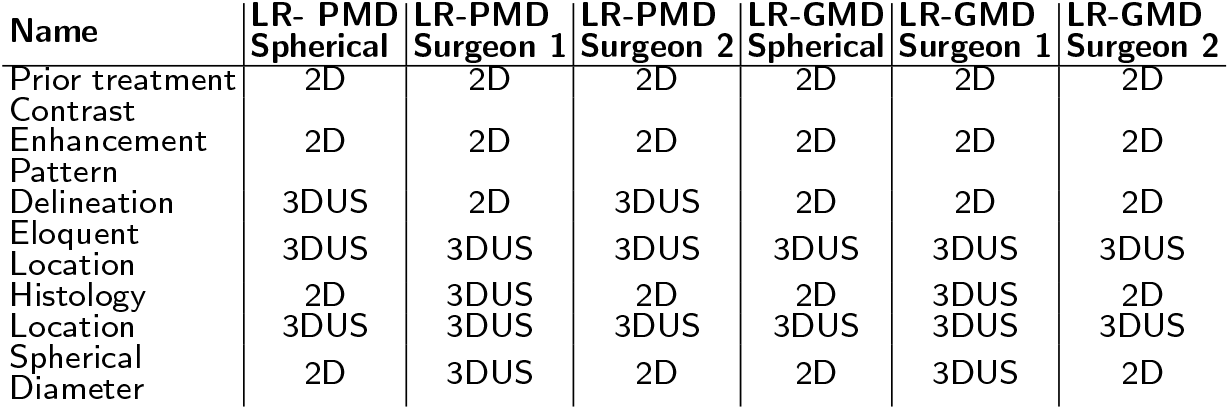
Likelihood of ultrasound of logistic regression based bagging models.

We have compared all logistic regression models using the chi-square test as depicted in Table 6. This table summarizes the important models that lead to our conclusions.

**Table 6:**
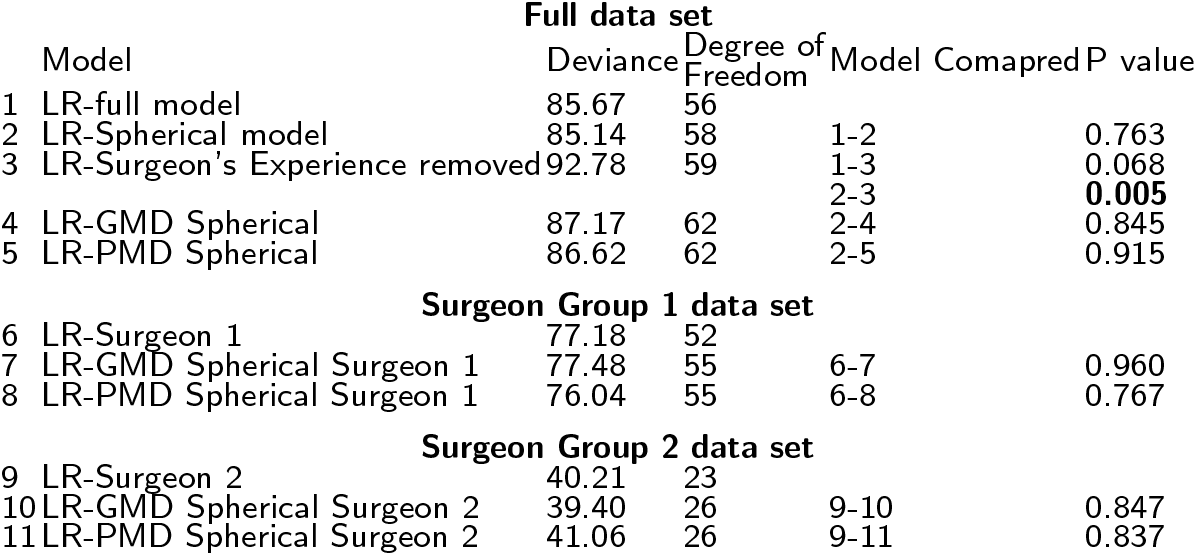
Statistical Analysis of different Models

Our composite models with redesigned variables showed performance comparable to the LR-spherical model. Hence, a decision may be taken with fewer parameters instead of all patient and tumor characteristics. This can also be concluded from the surgeon group 1 and surgeon group 2 data.

Table (7) shows the performance of all the models. RF-Surgeon 1 model resulted in all performance measures (accuracy, AUC ROC score, and AUC PR being more than 80%. All except LR-Surgeon Experience removed, and RF-Surgeon Experience removed models resulted in the AUC ROC score of more than 70%. The AUC ROC score 0.7 to 0.8 is considered acceptable, 0.8 to 0.9 is considered excellent, and more than 0.9 is considered outstanding [22]. Hence all our models expect LR-Surgeon’s Experience removed and RF-Surgeon’s Experience removed are acceptable.

**Table 7:**
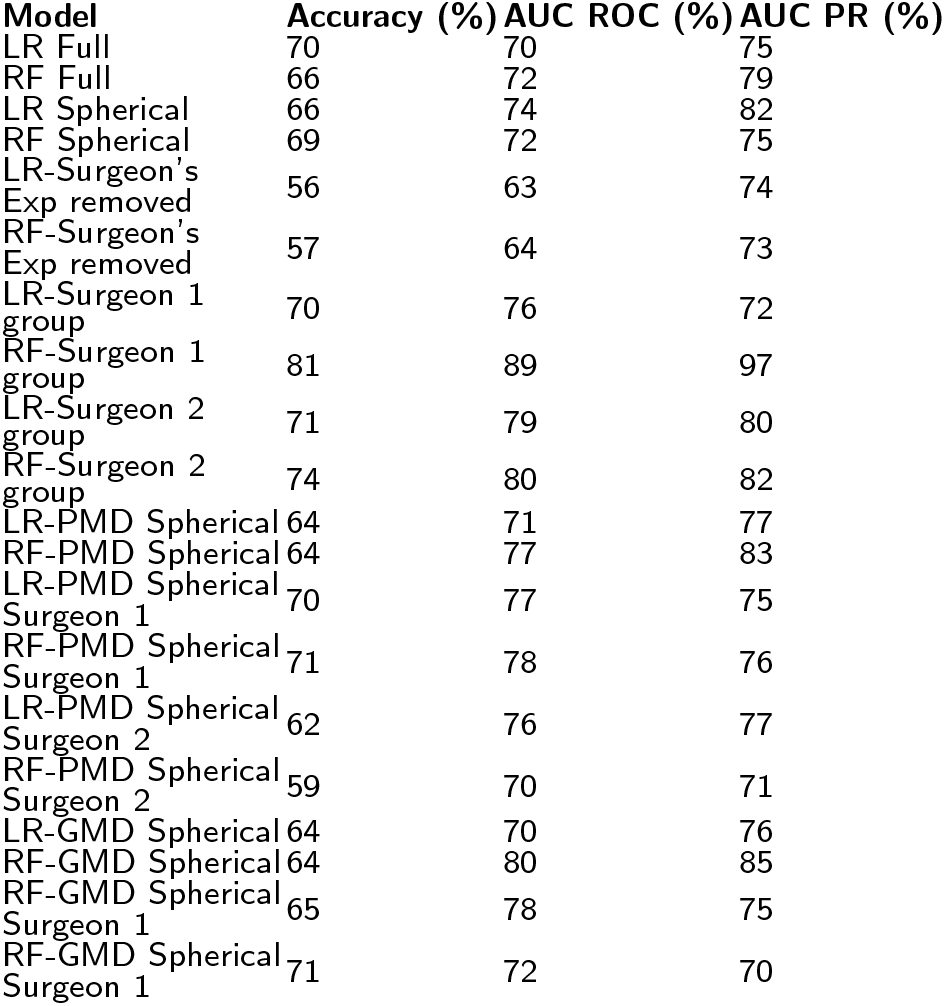
Performance of all models. AUC ROC denotes area under the receiver operating characteristic curve whereas AUC PR denotes area under the precisionrecall curve.

| Model | Accuracy (%) | AUC ROC (%) | AUC PR (%) |
| --- | --- | --- | --- |
| LR Full | 70 | 70 | 75 |
| RF Full | 66 | 72 | 79 |
| LR Spherical | 66 | 74 | 82 |
| RF Spherical | 69 | 72 | 75 |
| LR-Surgeon's<br>Exp removed | 56 | 63 | 74 |
| RF-Surgeon's<br>Exp removed | 57 | 64 | 73 |
| LR-Surgeon 1<br>group | 70 | 76 | 72 |
| RF-Surgeon 1<br>group | 81 | 89 | 97 |
| LR-Surgeon 2<br>group | 71 | 79 | 80 |
| RF-Surgeon 2<br>group | 74 | 80 | 82 |
| LR-PMD Spherical | 64 | 71 | 77 |
| RF-PMD Spherical | 64 | 77 | 83 |
| LR-PMD Spherical<br>Surgeon 1 | 70 | 77 | 75 |
| RF-PMD Spherical<br>Surgeon 1 | 71 | 78 | 76 |
| LR-PMD Spherical<br>Surgeon 2 | 62 | 76 | 77 |
| RF-PMD Spherical<br>Surgeon 2 | 59 | 70 | 71 |
| LR-GMD Spherical | 64 | 70 | 76 |
| RF-GMD Spherical | 64 | 80 | 85 |
| RF-GMD Spherical<br>Surgeon 1 | 65 | 78 | 75 |
| RF-GMD Spherical<br>Surgeon 1 | 71 | 72 | 70 |

## Discussion

1. All models discussed in the subsection () except LR-full and LR-Surgeon 1 group model favoring the navigated 3D ultrasound more likely when a patient is older.
2. All the logistic regression models discussed in subsection () agree that choice of 2D ultrasound is more likely when prior treatment is ‘yes’ or contrast enhancement pattern is ‘enhancing’ as shown in Table (4 - 5). As discussed with medical practitioners, 2D ultrasound is used either to localize the tumor or for a confirmatory scan whenever any prior treatment is done. Also, ‘enhancing’ tumors are clearly visible to surgeons; therefore, 2D ultrasound is enough. Wherever the tumor is in eloquent areas or the location is deep, all models recommend using navigated 3D ultrasound as the surgeon’s focus is to prevent damage to eloquent areas while achieving maximal possible resection. In cases, 2D ultrasound may not provide sufficient information about deeply situated tumors, then navigated 3D ultrasound is preferred.
3. The surgeon 1 model elaborated in the subsection () suggests the use of navigated 3D ultrasound for large spherical diameter tumors. In contrast, all other models trained on PMD and GMD data sets suggest the use of 2D ultrasound.
4. Except LR-PMD surgeon 2 and LR-PMD spherical, all other models discussed in the subsection () suggest the use of 2D ultrasound whenever delineation is good.
5. LR-PMD Spherical and LR-GMD Spherical models discussed in the section () have the same sign coefficient except for the delineation, which is defined in various ways. The random forest trained on these data sets also exhibits a different order of feature importance. Therefore, how surgeons interpret the moderate delineation is also an essential factor in deciding the type of intraoperative ultrasound.
6. The random forest-based models discussed in subsubsection () and () resulted spherical diameter as a most important factor in the models. Also, it is observed that feature importance is distinct for different models, which may be due to the surgeon’s personal choice that they would have for 2D or navigated 3D ultrasound.
7. We have also applied Principal Component Analysis (PCA) on the data set discussed in subsection, and found that it does not provide us a model with fewer dimensions that could explain intraoperative ultrasound decisions. However, after redesigning a few ordinal variables without compromising explainability, a more compact model with fewer features was obtained. This perhaps overcomes the limitation of the Principal Component Analysis method.

## Conclusion

In this paper, we have attempted to examine the factors that could have influenced the choice of use of a particular intraoperative imaging adjunct (US) in a large series of patients consecutively treated at a reference neurosurgery centre. Different logistic regression and random forest-based bagging models were fitted over the various data sets generated from a data set of 350 patients and tested on the test data sets. We found that the surgeon experience, prior treatment, and contrast enhancement pattern are statistically significant in almost all logistic regression-based models. The models trained on the surgeon’s stratified data sets show that patients’ age and histology are also statistically significant.

The random forest-based bagging model also showed that the surgeon experience and patient’s age are the two most important factors. The spherical diameter of the tumor is the essential attribute after removing the surgeon experience parameter from the model. The random forest-based model trained on the surgeon’s stratified group where only tumor characteristics are considered, depicts the distinct order of feature importance.

Logistic regression-based model highlights that likelihood of ultrasound type depends on how the delineation is considered. Therefore, we can say that different surgeons give different weightage to various features while selecting the intraoperative ultrasound. The models trained on the surgeon’s stratified data sets show that the surgeon’s personal choice affects the overall decision of intraoperative ultrasound.

We have introduced spherical diameter as a single parameter instead of three MRI measured dimensions of tumor. We found that one parameter, i.e., spherical diameter, is enough to capture the information of all three dimensions of the tumor. Tumor characteristics (delineation/ prior treatment/ contrast enhancement pattern/ eloquent monitoring/ histology/ location) were found to be adequate to explain the decision irrespective of patient characteristics (age, gender), by and large. Only in one subset of data, age plays some role in the decision-making. The 2D ultrasound was used more likely for previously treated superficial and enhancing tumors situated in non-eloquent areas. The navigated 3D ultrasound was used for non-enhancing tumors situated in eloquent areas and deep inside the brain.

The limitation of our work is that the results reflect associations between tumor factors and the use of a particular US type, but this cannot be interpreted as recommendations for the use of such US type in those subsets. For that, outcome analysis and correlation are important. However, analyzing choice distribution is important to be able to account for surgeon choices in future outcome comparison studies.

## Data Availability

Data is available with the authors and can be made available on request with reasonable justification.

## Appendix

### A Correlation Matrices

**Table 8:** Correlation computed using different methods for categorical, ordinal, and continuous variables.

| Name | Gender | Surgeon Exp | Prior treatment | Contrast Enhancement pattern | Delineation | Eloquent | Histology | Depth of tumor | Length | Width | Height | Glioma Location |
| --- | --- | --- | --- | --- | --- | --- | --- | --- | --- | --- | --- | --- |
| Age | 0.035 | 0.09 | -0.22 | 0.164 | -0.108 | -0.01 | 0.28 | 0.027 | -0.126 | -0.188 | -0.126 | 0.262 |
| Gender |  | 0.05 | -0.04 | -0.001 | 0.00 | 0.01 | 0.00 | 0.00 | -0.017 | -0.039 | 0.013 | -0.019 |
| Surgeon Exp |  |  | -0.08 | -0.019 | -0.01 | 0.08 | 0.06 | -0.022 | -0.032 | -0.057 | 0.013 | 0.034 |
| Prior treatment |  |  |  | 0.0 | 0.00 | -0.03 | 0.00 | -0.002 | -0.048 | -0.035 | -0.04 | 0.006 |
| Contrast Enhancement pattern |  |  |  |  | 0.24 | 0.00 | 0.15 | -0.019 | -0.078 | 0.012 | -0.042 | -0.002 |
| Delineation |  |  |  |  |  | 0.0 | -0.08 | -0.152 | -0.016 | -0.002 | 0.007 | -0.002 |
| Eloquent |  |  |  |  |  |  | 0.0 | -0.002 | 0.007 | 0.087 | 0.08 | -0.019 |
| Histology |  |  |  |  |  |  |  | 0.006 | -0.079 | -0.078 | -0.092 | -0.005 |
| Depth of tumor |  |  |  |  |  |  |  |  | -0.116 | -0.137 | -0.159 | 0.002 |
| Length |  |  |  |  |  |  |  |  |  | **0.697** | **0.683** | 0.013 |
| Width |  |  |  |  |  |  |  |  |  |  | **0.667** | -0.102 |
| Height |  |  |  |  |  |  |  |  |  |  |  | -0.024 |

**Table 9:** Correlation matrix with spherical diameter.

| Name | Gender | Surgeon Exp | prior treatment | Contrast Enhancement pattern | Delineation | Eloquent | Histology | Depth of tumor | Spherical Diameter | Glioma Location |
| --- | --- | --- | --- | --- | --- | --- | --- | --- | --- | --- |
| Age | 0.035 | 0.09 | -0.22 | 0.164 | -0.108 | -0.01 | 0.28 | 0.027 | -0.158 | 0.262 |
| Gender |  | 0.05 | -0.04 | -0.001 | 0.00 | 0.01 | 0.00 | 0.00 | -0.017 | -0.019 |
| Surgeon Exp |  |  | -0.08 | -0.019 | -0.01 | 0.08 | 0.06 | -0.022 | -0.035 | 0.034 |
| prior treatment |  |  |  | 0.00 | 0.00 | -0.03 | 0.00 | -0.002 | -0.047 | 0.006 |
| Contrast Enhancement pattern |  |  |  |  | 0.24 | 0.00 | 0.15 | -0.019 | 0.039 | -0.002 |
| Delineation |  |  |  |  |  | 0.00 | -0.08 | -0.152 | -0.006 | -0.002 |
| Eloquent |  |  |  |  |  |  | 0.00 | -0.002 | 0.065 | -0.019 |
| Histology |  |  |  |  |  |  |  | 0.006 | -0.089 | -0.005 |
| Depth of tumor |  |  |  |  |  |  |  |  | -0.157 | 0.002 |
| Spherical Diameter |  |  |  |  |  |  |  |  |  | -0.041 |

### B Statistical Analysis of data set of refined variables

**Table 10:**
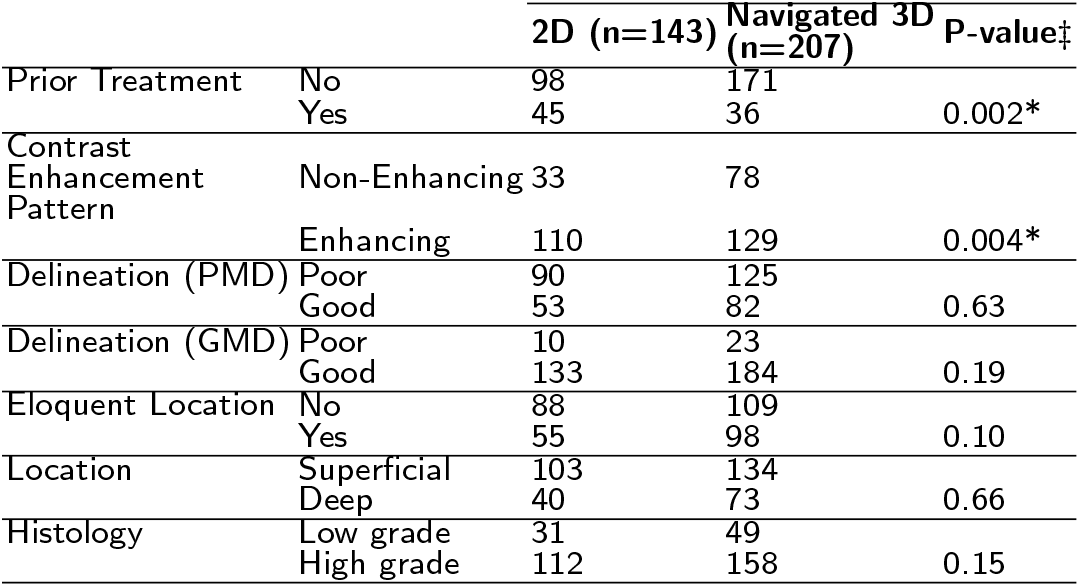
Distribution of data after converting ordinal variables to dichotomous. * denotes *p* < 0.05, p-values corresponding to Mann-Whitney test.

## Acknowledgements

None.

## Funding

Not applicable.

## Abbreviations

LR: Logistic Regression
RF: Random Forest

## Ethics approval and consent to participate

Ethics approval is obtained from the Institute Ethics committee. Consent of participants is not required as this is retrospective study.

## Competing interests

The authors declare that they have no competing interests.

## Consent for publication

All authors have given their consent for publication.

## Authors’ contributions

M.K. has performed the analysis and wrote the draft version of the manuscript. S.N., N.R., and A.M. contributed to the conceptualization of the problem and reviewed the draft version. A.M., P.S., and V.K.S. collected the data and helped in clinical inputs.

## Authors’ information

Text for this section…

## Notes

### Competing Interest Statement

The authors have declared no competing interest.

### Funding Statement

This study did not receive any funding.

### Author Declarations

Institutional Ethics Committee-III/900632 of Tata Memorial Centre gave ethics approval for this work

